# Reproductive health in Mexican women with systemic lupus erythematosus: pregnancy outcomes, menstrual irregularities and early menopause

**DOI:** 10.64898/2026.06.07.26354004

**Authors:** Sevilla-Parra Grecia, Bravo-García Fernanda, Mier y Terán Guevara MariaJose, Montes-Garcia Areli, Schäfer Alejandra, Rodríguez-Ochoa Nínive, Bienvenu Caballero Mercedes, Gonzalez Zenteno Said Gabriel, Peña-Ayala Angélica, Tinajero-Nieto Lizbet, Torres-Valdez Estefania, Martínez Domingo, Hernández Ledesma Ana Laura, Medina-Rivera Alejandra, Alpízar-Rodríguez Deshiré

## Abstract

**Objective:** To characterize pregnancy outcomes and menstrual irregularities in Mexican women with systemic lupus erythematosus (SLE) and identify clinical factors associated.

**Methods:** We conducted a cross-sectional study of women with SLE enrolled in the Mexican Lupus Registry (LupusRGMX) between May 2021 and September 2024. Clinical and reproductive data were collected using standardized questionnaires. Menopause was defined as the absence of menstruation for ≥12 consecutive months, and early menopause before age 40. Univariable and multivariable logistic regression analyses were used to identify factors associated with pregnancy complications and early menopause.

**Results:** A total of 210 women were included. Median age was 38 years (IQR 29–46) and median disease duration was 4 years (IQR 1–10). Among women with a history of pregnancy (47%), full-term delivery predominated (61%), while pregnancy loss occurred in 26% and preterm delivery in 13%. Pregnancy complications were reported in 9.6%, most commonly preeclampsia (6.7%). Younger maternal age was independently associated with pregnancy complications (OR 0.89, 95% CI 0.83–0.95) and adverse outcomes (OR 0.95, 95% CI 0.92–0.98). Higher disease activity was associated with complications in univariable analysis. Most pregnancies (68.3%) occurred before diagnosis. Early menopause was observed in 6.2% and was independently associated with longer disease duration.

**Conclusion:** Younger maternal age was independently associated with adverse pregnancy outcomes in women enrolled in the LupusRGMX. Most pregnancies occurred prior to SLE diagnosis. Early menopause was associated with longer disease duration, suggesting the impact of cumulative disease burden on ovarian function.

## 1 Introduction

Systemic Lupus Erythematosus (SLE) is a chronic, systemic, autoimmune disease characterized by a complex pathogenesis. The disease primarily affects women of childbearing age, with a female to male ratio of 9 to 1 (1). Given this disproportionate impact, SLE poses unique challenges to reproductive health, particularly in low and middle-income regions where access to specialized care may be limited (2–4).

Depending on the magnitude of the organ involvement and disease activity, ovarian dysfunction, menstrual irregularities, infertility, and poor obstetric outcomes occur more frequently among women with SLE (4–6). Even when conception is achieved, the elevated incidence of adverse pregnancy outcomes remains a major clinical challenge (1). Outcomes and complications of pregnancy are associated with disease activity (1,5). Preeclampsia/eclampsia, active SLE, and thrombocytopenia were strongly associated with preterm delivery and maternal SLE flares, making them major predictors of fetal death (4,5,7). In contrast, optimal timing of conception, careful monitoring of disease activity, multidisciplinary management approaches, and clinical advances have contributed to reductions in maternal morbidity and mortality, allowing most women of reproductive age with SLE to complete one or more pregnancies (8). Cytotoxic drugs used to treat SLE, such as cyclophosphamide, can decrease the ovarian reserve and may lead to premature ovarian failure (9).

Studies reported 11.7% of amenorrhea among juvenile SLE patients, with no significant differences observed between patients with and without amenorrhea for frequency, cumulative dose, number of pulses, or duration of intravenous cyclophosphamide therapy. In contrast, amenorrhea was significantly associated with higher disease activity and greater cumulative organ damage (10).

In Mexico, evidence from a study in women with autoimmune rheumatic diseases highlights substantial unmet needs in sexual and reproductive health (11). Patients report uncertainty regarding the safety and effectiveness of contraceptive methods in the context of their disease and treatment, as well as anxiety related to infertility, hereditary risk, and pregnancy outcomes, which complicates reproductive planning. Concerns about the safety of medication during pregnancy and breastfeeding indicate a lack of guidance and information from healthcare providers, and recognize the importance of coordinated and multidisciplinary care (11).

SLE significantly affects key domains of reproductive health, including fertility, pregnancy outcomes, and contraceptive decision-making, requiring individualized counseling and multidisciplinary care to optimize maternal and fetal outcomes (12). Although reproductive health outcomes in SLE have been studied in other populations, evidence from Mexican and broader Latin American populations remains limited (13,14). Differences in healthcare access, socioeconomic conditions, and cultural context may influence reproductive health experiences and outcomes, underscoring the need for patient-reported data from Mexican women with SLE to inform culturally and contextually appropriate care strategies (15). This study aims to address this gap by exploring pregnancy outcomes and menstrual irregularities in Mexican people with SLE and identify clinical factors associated.

## 2 Materials and methods

### 2.1 Study design and population

We conducted a comparative cross-sectional study. The study protocol was reviewed and approved by the Research Ethics Committee of the Instituto de Neurobiología at the Universidad Nacional Autónoma de México (UNAM, approval no. 093.H) on September 8, 2025. All collected data were anonymized and securely stored at the National Laboratory of Advanced Scientific Visualization (LAVIS) at UNAM. Participants provided written informed consent and received a copy of the privacy notice in accordance with the Mexican Federal Law on the Protection of Personal Data Held by Private Parties.

Participants were recruited between May 2021 and September 2024 through two complementary strategies: referral by rheumatologists affiliated with the Mexican Lupus Registry (LupusRGMX) and an open call disseminated through the registry’s online platform (16). Details regarding the LupusRGMX cohort have been reported previously (16,17).

Eligibility required a confirmed SLE diagnosis based on the American College of Rheumatology and the Systemic Lupus International Collaborating Clinics (ACR/SLICC) classification criteria (18). Controls included individuals without a self-reported SLE diagnosis from the LupusRGMX database, as well as Mexican volunteers from the Jaguar Project (https://jaguar.liigh.unam.mx/). For the control group individuals reporting systemic autoimmune, rheumatologic conditions, or a history of organ transplantation, were excluded (see Supplementary Table 1). For both the SLE and the control group, only individuals aged 18 years or older were included. Records with incomplete information were excluded.

### 2.2 Reproductive health questionnaire

The reproductive health questionnaire was developed based on items from the 2021 questionnaire of the Nurses’ Health Study (19). The items included were adapted to capture information on menstrual history, reproductive events, and gynecologic health while maintaining consistency with the structure and content of the original instrument.

### 2.3 Clinical variables

Clinical variables were collected through standardized questionnaires from the LupusRGMX. All variables were self-reported, unless otherwise specified.

The interval between symptom onset and SLE diagnosis was calculated using predefined categorical intervals ranging from <6 months to ≥10 years prior to diagnosis. The highest category (≥10 years) represents an open-ended interval and may include participants with substantially longer pre-diagnostic periods.

Pregnancy history was defined as the report of at least one prior pregnancy. Adverse pregnancy outcomes were defined as a composite outcome including pregnancy loss, preterm delivery, or the occurrence of pregnancy-related complications. Pregnancy complications were assessed at the pregnancy level and defined as the occurrence of preeclampsia, eclampsia, or HELLP syndrome. Preterm delivery was defined as delivery occurring before 37 weeks of gestation. Pregnancy loss was defined as self-reported miscarriage or stillbirth. The number of pregnancies was treated as a discrete variable based on total pregnancies.

Menopause was defined as the absence of menstruation or amenorrhea for at least 12 consecutive months that reflects ovarian dysfunction (20). For participants meeting this criterion, the reported age at the last menstrual period was verified to be earlier than the age at the time of data collection. Early menopause was defined as menopause onset before 40 years of age. Surgically induced menopause was defined as the history of hysterectomy and/or oophorectomy. Reproductive lifespan was calculated as the difference between age at menarche and age at the last menstrual period.

Disease activity was assessed using the SLEDAI index, with active disease defined as a score >6. Cumulative organ damage was evaluated using the SLICC damage index. Lupus nephritis was defined based on prior clinical diagnosis. Treatment at the time of registry inclusion was recorded. Information on cumulative dose, duration of therapy, and prior treatment exposure was not available and therefore, was not included in the analysis.

### 2.4 Statistical analysis

Statistical analyses were performed using R software (version 4.4.2). Continuous variables were summarized as median and interquartile range (IQR), and categorical variables as frequencies and percentages. Analyses were conducted at both the individual level (per participant) and the pregnancy level (per reported pregnancy), depending on the outcome of interest.

Group comparisons were conducted using non-parametric tests due to the non-normal distribution of continuous variables. The Wilcoxon rank-sum test was used for comparisons between two groups, and the Kruskal–Wallis test was applied for comparisons across three or more groups. Categorical variables were compared using Pearson’s chi-square test or Fisher’s exact test, as appropriate.

Associations between the number of pregnancies and clinical variables were evaluated using Spearman’s rank correlation coefficients. Multivariable logistic regression models were constructed to identify factors independently associated with pregnancy complications and adverse pregnancy outcomes. Covariates were selected *a priori* based on clinical relevance and prior literature, with consideration of key variables, such as age, reproductive lifespan, disease activity (SLEDAI), disease duration, nephritis, and treatment exposure. Model estimates were reported as odds ratios (ORs) with 95% confidence intervals (CIs).

To evaluate factors associated with early menopause, a penalized logistic regression model using Firth’s method was fitted to account for the limited number of events and to reduce potential small-sample bias. Predicted probabilities were derived from the fitted model to estimate the relationship between disease duration and early menopause while adjusting for age. All statistical tests were two-sided, and a p-value <0.05 was considered statistically significant. The code used to perform the analyses and generate the figures is publicly available at https://github.com/NeuroGenomicsMX/Reproductive_Health_SLE

## 3 Results

### 3.1 Study population and reproductive profile in women with SLE

A total of 234 records were identified from the Mexican Lupus Registry database. After applying the inclusion criteria and verifying data completeness, 210 women were included (Figure 1). To explore potential differences between women with SLE (n=210) and non-SLE affected controls (n=112), we performed an exploratory comparative analysis (Supplementary Table 2). Baseline characteristics of SLE patients are shown in Table 1. Women with SLE were older than controls (median age 38 vs 26 years), limiting direct comparisons. Age at menarche was similar between groups. Although women with SLE appeared to have a lower frequency of pregnancy history and a higher proportion of reported pregnancy complications compared with controls, these findings should be interpreted with caution, given the differences in age distribution and reproductive exposure between groups.

**Figure 1.**
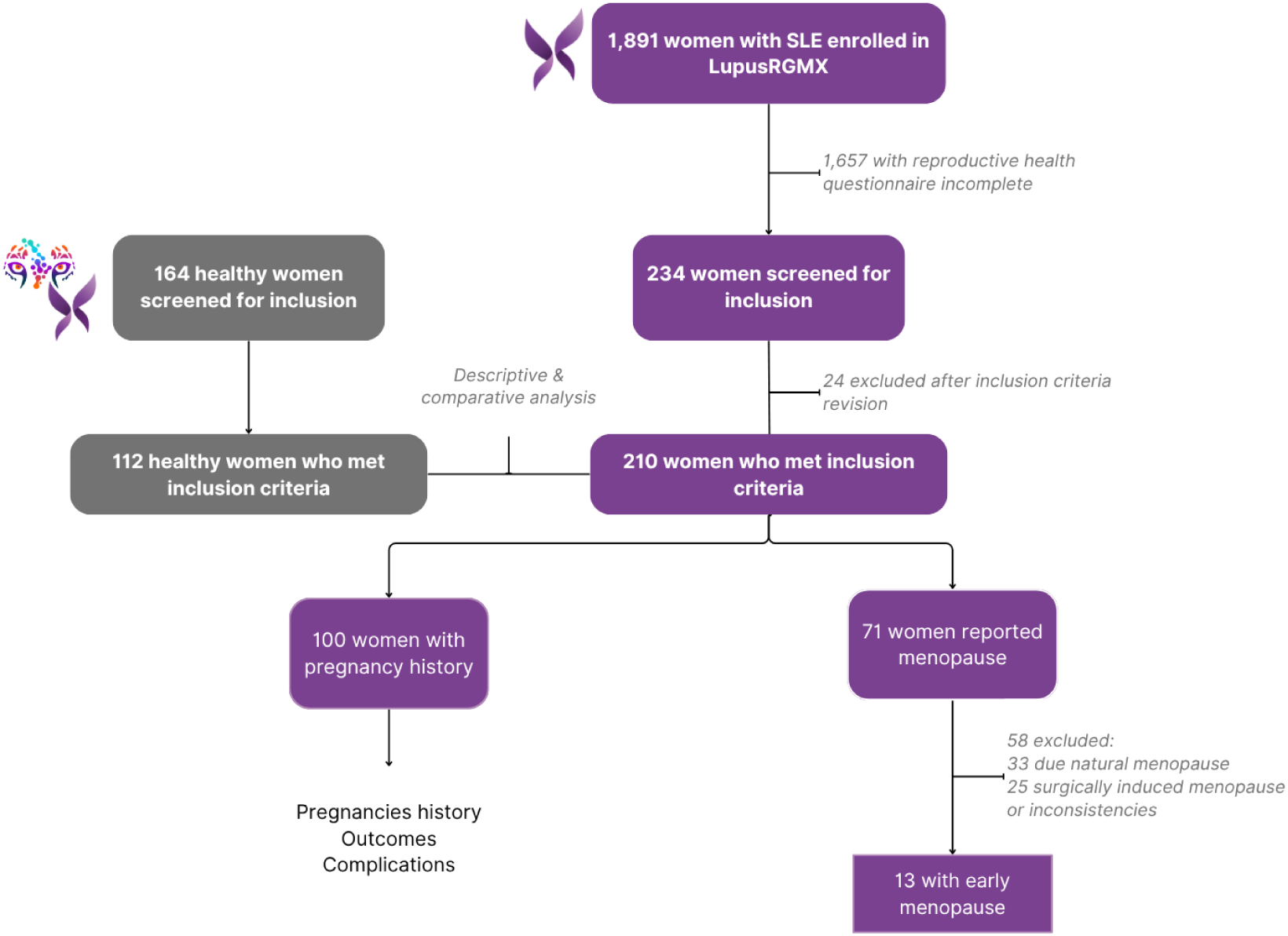
Study flow diagram. Participants with systemic lupus erythematosus (SLE) are shown in purple, while participants without SLE (control group) are shown in gray.

**Table 1.** Demographic, clinical, and reproductive characteristics of women with SLE (n = 210).

| Variable | SLE<br>n = 210<sup>1</sup> |
| --- | --- |
| **DEMOGRAPHIC** |  |
| Age (yrs) | 38 (29, 46) |

| **REPRODUCTIVE CHARACTERISTICS** |  |
| --- | --- |
| **Age at menarche (yrs)** | 12 (11, 13) |
| **Last period age (yrs)** | 35 (29, 44) |
| **Reproductive life (yrs)** | 24 (16, 31) |
| **Menstrual cycle regularity** |  |
| Yes | 139 (66%) |
| No | 68 (32%) |
| Unknown | 3 (1.4%) |
| **Pregnancy history** |  |
| Currently pregnant | 0 (0%) |
| History of pregnancy | 100 (47%) |
| Pregnancy complications | 20/100 (20%) |
| Number of pregnancies |  |
| 1 | 33 (33%) |
| 2 | 35 (35%) |
| 3 | 24 (24%) |
| 4 | 7 (7%) |
| 5 | 1 (1%) |
| Pregnancy loss | 32/100 (32%) |
| Gestational diabetes | 3/100 (3%) |
| Gestational hypertension | 14/100 (14%) |
| Ectopic pregnancy | 5/100 (5%) |
| **Gynecologic surgical history** |  |
| Hysterectomy | 24 (11%) |
| Oophorectomy | 17 (8.1%) |
| **Polyendocrine Metabolic Ovarian Syndrome (PMOS)** | 30 (14%) |
| **Use of contraceptives** | 22 (10%) |
| **Natural menopause**<sup>3</sup> | 33 (15.7%) |
| **Early menopause** | 13 (6.2%) |
| **Age at natural menopause** | 48 (45, 50) |
| <sup>1</sup> Median (IQR) or Frequency (%) |  |
| <sup>2</sup> Kruskal-Wallis rank sum test; Fisher's exact test; Pearson's Chi-squared test |  |
| <sup>3</sup> Excluding hysterectomy and/or oophorectomy |  |

To further characterize reproductive health-related features, subsequent analyses focused exclusively on women with SLE (Table 1). The cohort had a median disease duration of 4 years (IQR 1–10), consistent with an early-stage disease population. Disease activity was moderate to high, with a median SLEDAI score of 21 (IQR 12–32), while cumulative organ damage was low to moderate, as reflected by a median SLICC damage index of 2.0 (IQR 1.0–4.0). Renal involvement was present in 19.5% of patients. Most participants were receiving antimalarials (75.7%) and corticosteroids (71.4%). Participants were recruited from multiple regions across Mexico, reflecting broad geographic representation. The cohort included women across a wide age range, with the highest proportion observed among those aged 18 to 30 years (Supplementary Figure 1).

### 3.2 Pregnancy outcomes in SLE

Among the participants, 100 (47%) reported at least one pregnancy, accounting for a total of 208 pregnancies (Table 1; Supplementary Table 3). When pregnancies were stratified according to number (1, 2, or ≥3), women with a higher number of pregnancies were older (p=0.044) (Supplementary Table 3). No significant differences were observed across pregnancy number in disease activity (SLEDAI score), nephritis prevalence, or treatment exposure. Pregnancy outcomes and complication rates did not differ significantly.

Figure 2 illustrates the distribution of pregnancy outcomes and their relationship with obstetric complications. Full-term deliveries predominated, whereas pregnancy loss and preterm delivery were less frequent (Figure 2A). The Sankey diagram highlights the flow between pregnancy number, outcomes, and complications, revealing that severe complications such as preeclampsia and HELLP syndrome were more commonly associated with preterm deliveries.

**Figure 2.**
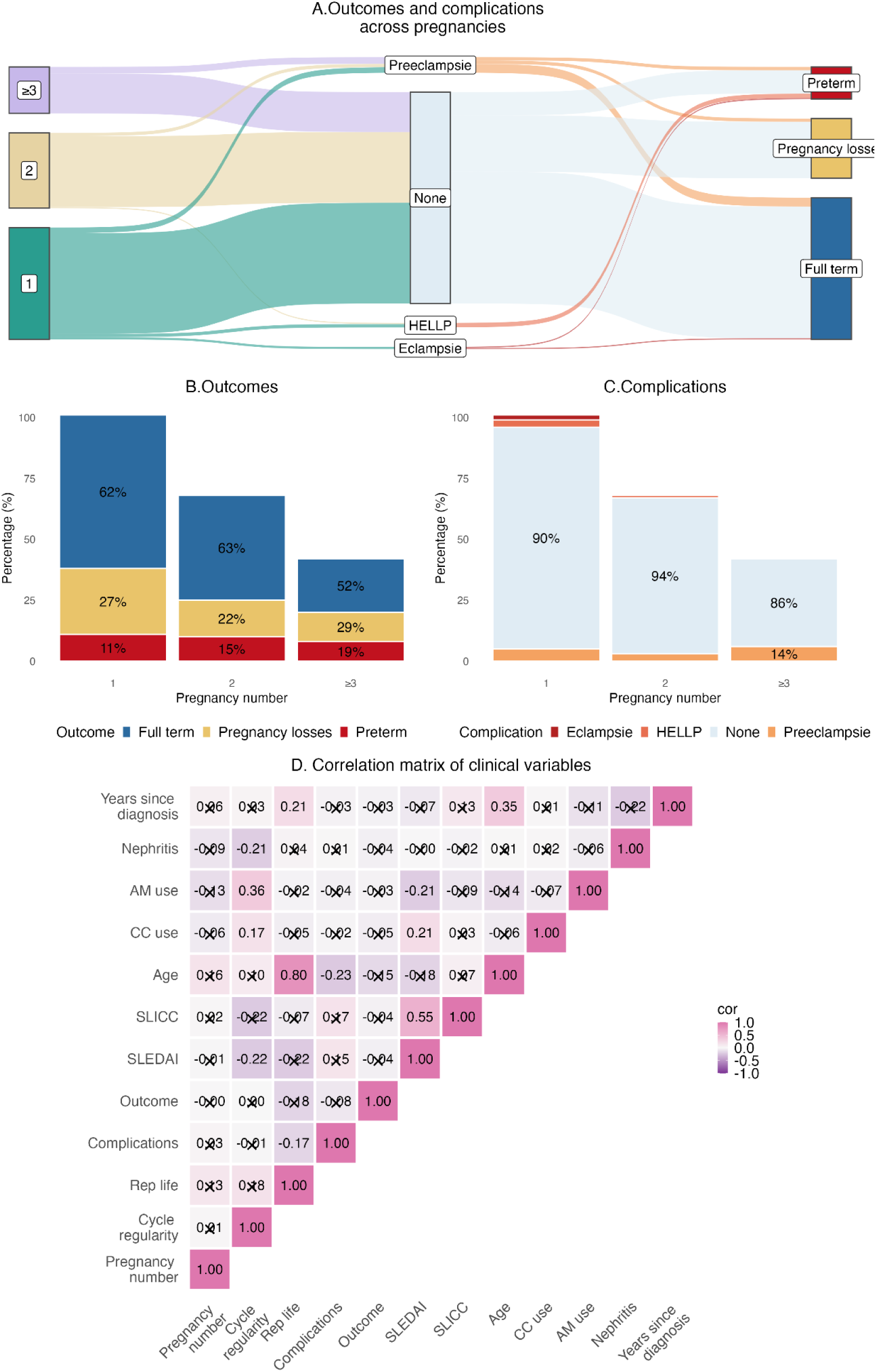
Distribution of pregnancy outcomes, complications, and their relationship with clinical variables in women with SLE. (A) Sankey diagram illustrating the flow between pregnancy number, pregnancy outcomes, and obstetric complications. Colors represent pregnancy outcomes (blue: full term; yellow: pregnancy loss; red: preterm) and complications (light blue-gray, no complications; orange, preeclampsia; salmon, HELLP; dark red, eclampsia). (B) Distribution of pregnancy outcomes according to pregnancy number, showing the predominance of full-term deliveries. Bars are color-coded as follows: blue, full term; yellow, pregnancy loss; red, preterm. (C) Frequency of pregnancy-related complications across pregnancy groups, highlighting the overall low prevalence of adverse events. Bars are color-coded as follows: light blue-gray, no complications; orange, preeclampsia; salmon, HELLP; dark red, eclampsia. (D) Spearman correlation matrix of pregnancy number and clinical variables. Cell color and numeric values represent correlation coefficients (r), while non-significant associations (adjusted p > 0.05, Benjamini–Hochberg) are indicated by an ‘×’.

Pregnancy outcomes were predominantly favorable, with full-term delivery occurring in 61% of pregnancies (Figure 2B). Pregnancy loss was reported in 26% of pregnancies and preterm delivery in 13%, while the overall prevalence of pregnancy-related complications remained low (9.6%), including preeclampsia (6.7%), HELLP syndrome (1.9%), and eclampsia (1.0%). No significant differences in disease activity, nephritis, or treatment exposure were observed across pregnancy order (Figure 2C). Consistent with these findings, Spearman correlation analysis showed a strong positive correlation between age and reproductive life (r = 0.80, padj < 0.0001), whereas no significant correlations were identified between pregnancy number and disease activity (SLEDAI), cumulative damage (SLICC), nephritis, or treatment use (Figure 2D).

To evaluate clinical and treatment factors associated with pregnancy outcomes, comparative analyses were performed across outcomes: pregnancy loss, full-term delivery, and preterm delivery (Supplementary Table 4). Maternal age differed significantly (p=0.032), as did reproductive life (p=0.006), with longer reproductive life duration observed among full-term pregnancies. In contrast, no significant differences were observed in disease activity, nephritis prevalence, or treatment exposure across. Severe obstetric complications were disproportionately represented among preterm pregnancies (p<0.001), whereas the majority of full-term and pregnancy loss cases occurred in the absence of complications.

To investigate factors associated with the development of pregnancy complications, comparative analyses were performed (Supplementary Table 5). Pregnancies complicated by adverse events occurred in younger women (p<0.001) and were associated with shorter reproductive life (p=0.009). In addition, higher disease activity was observed among pregnancies with complications compared to those without (p=0.021). No significant differences were observed in nephritis prevalence or treatment exposure.

In multivariable logistic regression analyses (Supplementary Table 6), age emerged as the only independent factor associated with pregnancy complications (OR 0.89, 95% CI 0.83–0.95) and adverse outcomes (OR 0.95, 95% CI 0.92–0.98). Age at menarche showed a borderline inverse association with complications (OR 0.70, 95% CI 0.47–0.98, p=0.051).

In the multivariable analysis (Figure 3), maternal age was the only variable independently associated with pregnancy complications (OR ≈ 0.87). Disease activity, cumulative damage, reproductive life, disease duration, and treatment exposure were not significantly associated. Likewise, no independent predictors of pregnancy outcomes were identified, although antimalarial use showed a non-significant positive association (OR ≈ 1.95).

**Figure 3.**
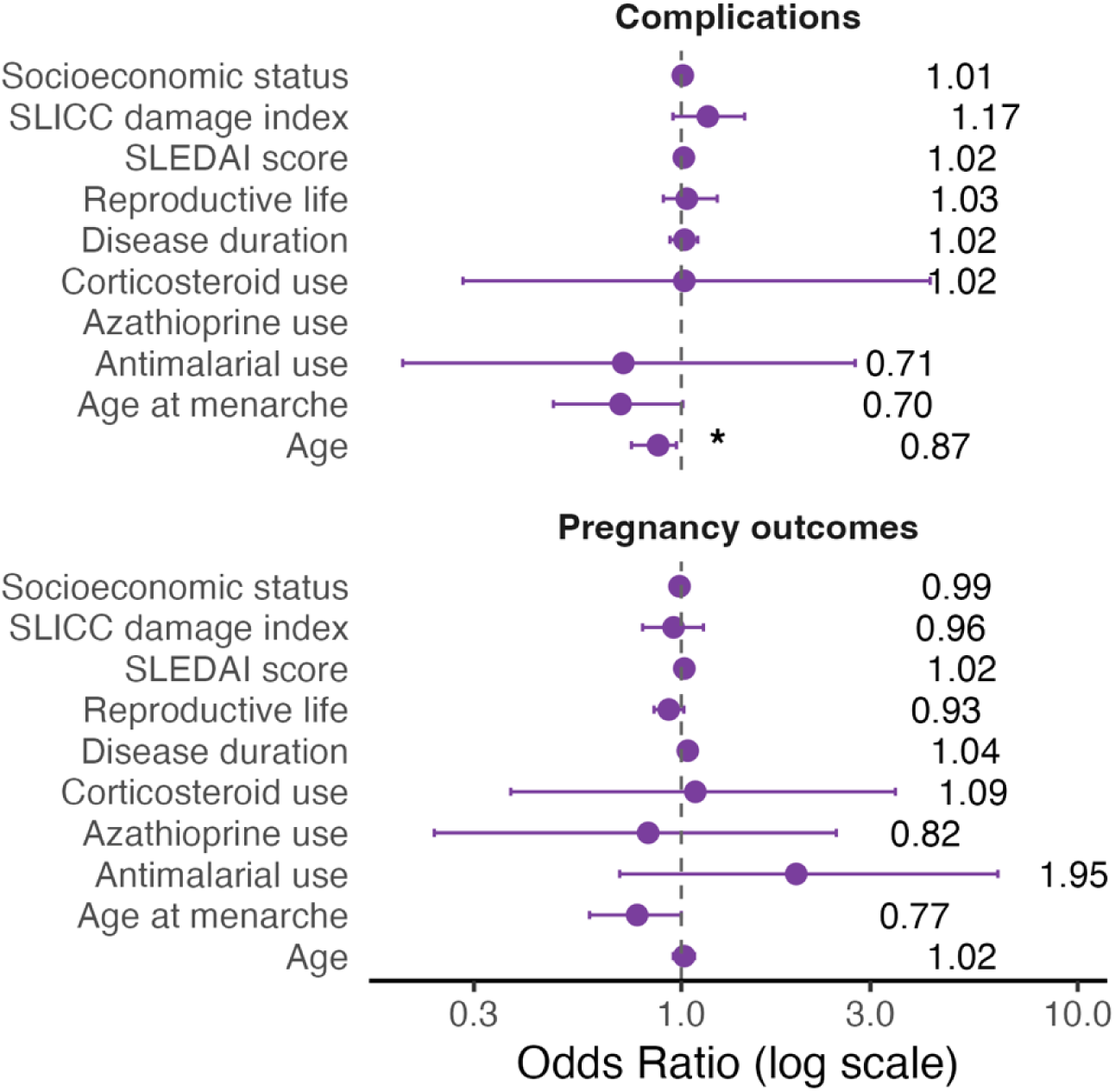
Multivariable logistic regression analysis of factors associated with pregnancy complications (top) and adverse pregnancy outcomes (bottom) in women with SLE. Each point represents the estimated odds ratio (OR), and horizontal lines indicate the 95% confidence intervals. The vertical dashed line at OR = 1 represents the null effect; estimates whose confidence intervals cross this line are not statistically significant.

The temporal distribution of pregnancies relative to SLE diagnosis is shown in Figure 4. Each horizontal trajectory represents the reproductive timeline of a participant, including the onset of lupus-associated symptoms, pregnancies, and their outcomes. To characterize temporal patterns associated with adverse outcomes, only participants who experienced pregnancy complications and/or pregnancy loss were included.

**Figure 4.**
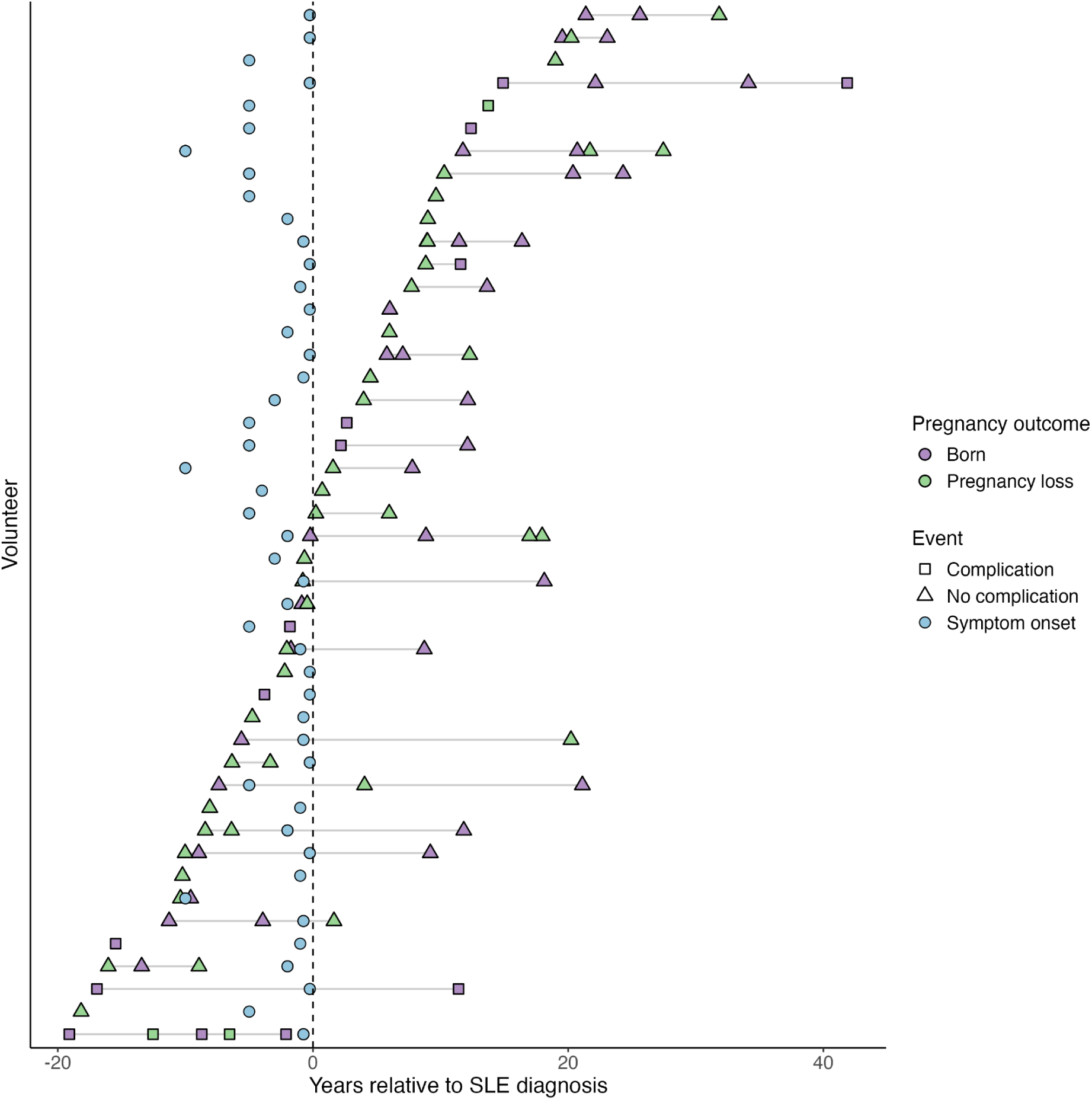
Individual timelines of symptom onset and pregnancy events relative to SLE diagnosis. Volunteers who experienced pregnancy complications and/or pregnancy loss were included in this analysis; only those with available information on symptom onset were considered (n = 94). Each horizontal line represents a participant with SLE. Points indicate pregnancies occurring before or after SLE diagnosis, expressed as years relative to diagnosis (dashed vertical line, time = 0). Point colors represent pregnancy outcomes (purple: live birth; green: pregnancy loss), and shapes indicate the presence or absence of pregnancy complications. Blue circles denote the reported onset of SLE-associated symptoms prior to diagnosis.

Notably, only 48 pregnancies occurred after diagnosis. Symptom onset frequently preceded formal diagnosis by several years, consistent with the documented delay between initial manifestations and clinical recognition of SLE. A complete overview of reproductive timelines for the full cohort is provided in Supplementary Figure S2.

Most pregnancies occurred prior to SLE diagnosis (68.3%). Both live births and pregnancy losses were observed throughout the reproductive timeline, and complications occurred in both pre and post diagnosis periods. Importantly, when integrating these temporal patterns with the comparative analyses, pregnancy complications were more frequent among younger women and those with higher disease activity (Supplementary Table 5).

Although most pregnancies resulted in favorable outcomes, variability in maternal age and disease activity underscores the need to consider reproductive health beyond pregnancy-related events. Therefore, we evaluated long-term ovarian function by assessing the prevalence and clinical correlates of menopause and early menopause.

### 3.3 Factors associated with early menopause in women with systemic lupus erythematosus

We evaluated the prevalence of menopause and early menopause and explored their associations with clinical and reproductive variables. After excluding women with surgically induced menopause, 183 participants were included (Supplementary Table 7). Early menopause was observed in 13 women (6%).

Women with menopause were older and had longer disease duration compared with menstruating women (both p<0.001). Age at the last menstrual period also differed significantly (p<0.001). Pregnancy history was more frequent among women with menopause (p<0.001), and a modest difference in age at menarche was observed (p=0.012).

Use of antimalarials was less frequent among women with menopause and early menopause compared with menstruating women (p<0.001). In contrast, no significant differences were observed in disease activity, cumulative organ damage, corticosteroid exposure or dose, immunosuppressive therapy, nephritis prevalence, smoking status, body mass index, or socioeconomic status.

Women with menopause had a longer disease duration than menstruating women (Figure 5A). In penalized logistic regression analysis, both longer disease duration (OR = 1.08, 95% CI: 1.01–1.16) and older age (OR = 1.08, 95% CI: 1.01–1.18) were independently associated with higher odds of early menopause (Figure 5B). Consistent with these findings, the age-adjusted predicted probability of early menopause increased progressively with disease duration (Figure 5C).

**Figure 5.**
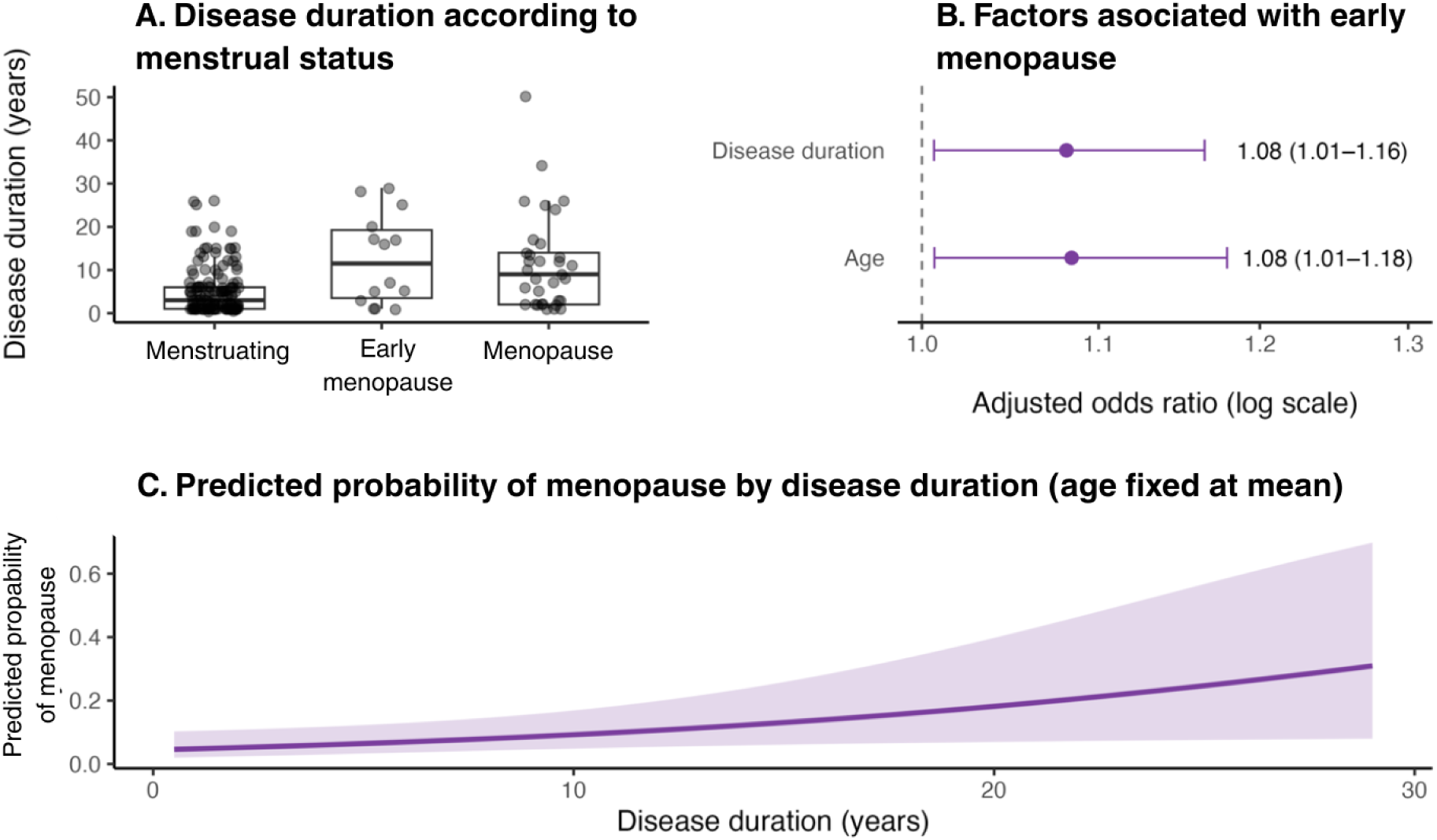
Early menopause in women with SLE. (A) Disease duration according to menstrual status. (B) Penalized logistic regression analysis showing adjusted odds ratios for early menopause. (C) Predicted probability of early menopause according to disease duration, adjusted for age.

## 4 Discussion

In this study, we evaluated reproductive health outcomes, including pregnancy history, pregnancy complications, and reproductive lifespan of women with SLE. We found that younger maternal age was independently associated with pregnancy complications, while disease activity showed association only in univariable analyses. Longer disease duration was associated with an increased likelihood of early menopause. Whereas, nephritis and treatment exposure were not significantly associated with adverse pregnancy outcomes. Together, these findings highlight the complex relationship between reproductive health and disease-associated factors in women with SLE.

Previous studies have shown that women with SLE are at increased risk of adverse obstetric outcomes, including stillbirth, preeclampsia, preterm delivery, and intrauterine growth restriction (4,21,22). These complications have been associated with thrombocytopenia, hypocomplementemia, proteinuria, higher disease activity, and high-dose corticosteroid use (22). In a Mexican cohort, active disease before pregnancy was a strong predictor of maternal complications, whereas antimalarial use was protective (23). Similarly, proteinuria in early pregnancy and preeclampsia were identified as major risk factors for adverse fetal outcomes, highlighting the central role of disease activity in determining pregnancy outcomes (24).

Interpreting reproductive outcomes women with SLE requires considering the limited availability of population-based data on reproductive health indicators in the country. Most published evidence derives from specific cohorts rather than nationally representative samples. For example, the Mexican Teachers’ Cohort has provided important information on reproductive history and hormonal factors among Mexican women, although it represents a specific occupational group (25). Earlier work conducted among female workers affiliated with the Instituto Mexicano del Seguro Social also noted the scarcity of population-based data on reproductive aging and menopause in Mexico (26). Additionally, studies in Mexican women with autoimmune rheumatic diseases have shown suboptimal contraceptive use and reproductive counseling, particularly among those receiving teratogenic therapies, highlighting persistent gaps in reproductive health education and multidisciplinary care (27).

Our findings indicate that pregnancy outcomes in women with SLE were generally favorable, with a predominance of full-term deliveries and a relatively low frequency of complications (Figure 3). However, important patterns emerged when exploring factors associated with adverse outcomes and their temporal relationship with disease onset and diagnosis.

Previous studies have identified disease activity as one of the strongest predictors of adverse pregnancy outcomes in SLE (21). In a prospective Mexican cohort of SLE pregnancies, disease activity both before and during pregnancy was independently associated with the development of preeclampsia, which in turn was linked to prematurity, low birth weight, and neonatal mortality. Notably, antimalarial use during pregnancy was associated with a reduced risk of preeclampsia (28). Consistent with these findings, higher disease activity in our cohort was significantly associated with pregnancy complications despite the overall favorable pregnancy outcomes (Supplementary Table 6).

Interestingly, development of pregnancy complications occurred in younger women (Figure 4). While age is often considered a risk factor in the general obstetric population, this finding may reflect a different underlying mechanism in SLE. Younger age at pregnancy could be associated with earlier or more aggressive disease phenotypes, or with pregnancies occurring closer to the onset of disease activity, when immune dysregulation is not yet adequately controlled (22,29,30).

The timeline analysis of the pregnancies and SLE onset and diagnosis (Figure 5) provides additional insight into this observation. Several pregnancies occurred *prior* to the formal diagnosis of SLE, and symptom onset frequently preceded diagnosis by several years. This suggests that a substantial proportion of pregnancies may have taken place during a preclinical or undiagnosed phase. Given that SLE-associated immune alterations can be present before diagnosis, it is plausible that subclinical disease activity may have contributed to adverse pregnancy outcomes, even in the absence of a formal SLE diagnosis (31,32). Importantly, pregnancy complications were observed both before and after diagnosis, indicating that risk is not exclusive of post-diagnosis pregnancies. Instead, these findings support a continuum model in which disease activity, rather than the timing of diagnosis, plays a central role in shaping reproductive outcomes.

Our findings support *prior* evidence by highlighting the role of disease activity and age, contrary to expectations, pregnancy complications were not associated with nephritis or treatment exposure in our cohort. This may be partly explained by the low prevalence of nephritis in our cohort (16%), which limited statistical power, and by the lack of detailed treatment data (duration, dosage, and adherence), which constrained the evaluation of treatment-related effects (21).

Beyond pregnancy outcomes, our findings also highlight the importance of long-term ovarian function as a key component of reproductive health in women with SLE. Current evidence suggests that SLE *per se* does not necessarily impair baseline fertility; however, high disease activity and exposure to cytotoxic therapies can adversely affect reproductive function (33–35). Reported complications include secondary amenorrhea during severe disease flares, hypofertility associated with renal insufficiency, and ovarian insufficiency related to cyclophosphamide (CTX) exposure (34–38).

In this study, early menopause was associated with longer disease duration, suggesting that cumulative disease burden may play a role in ovarian dysfunction. This finding is consistent with reports indicating an increased prevalence of premature ovarian insufficiency and earlier menopause among women with SLE (39,40). The mechanisms underlying ovarian dysfunction in SLE are likely multifactorial, involving chronic immune activation, autoantibody-mediated ovarian damage, prolonged systemic inflammation, and exposure to gonadotoxic therapies, particularly cyclophosphamide (CTX) (34,35,41,42). However, evidence regarding the relative contribution of disease versus treatment-related factors remains inconsistent. Large cohort studies have shown that, in the absence of CTX exposure, the prevalence of premature ovarian failure and the median age at natural menopause are similar to those observed in the general population, highlighting the heterogeneity of reproductive outcomes in SLE (43,44). Likewise, treatment exposure was not significantly associated with early menopause in our cohort, although this result should be interpreted cautiously given the very small number of patients exposed to CTX (n=1), which limited our ability to detect such an association.

Interestingly, disease activity was not significantly associated with early menopause, in contrast to its role in pregnancy complications. This may reflect differences in the temporal dynamics of these outcomes, where pregnancy complications are influenced by short-term fluctuations in disease activity, whereas amenorrhea or menopause may represent the cumulative effect of chronic disease exposure over time. The association between disease duration and early menopause reinforces the need to consider reproductive aging and fertility preservation as part of the longitudinal management of women with SLE.

In this cohort, reproductive outcomes were heterogeneous yet clinically meaningful. While most pregnancies resulted in favorable outcomes, maternal age was independently associated with complications, underscoring the importance of individualized risk assessment and optimal disease control before and during pregnancy. The observed prevalence of menopause, particularly its early onset, and its association with longer disease duration suggest that cumulative disease exposure may influence long-term reproductive trajectories. Together, these findings support a longitudinal approach to reproductive health in SLE, not limited to pregnancy. Systematic assessment of menstrual history, fertility intentions, and ovarian function should be integrated into routine clinical care to inform decision-making and improve patient-centered outcomes.

Several limitations should be considered when interpreting the findings from comparisons between SLE and controls. Women with SLE were older than controls, which likely reflects differences in cumulative reproductive exposure and limits direct comparisons of reproductive outcomes between groups. Ideally, analyses would include age-matched population controls; however, comparable datasets for Mexican women are scarce and not readily accessible. The Jaguar control group therefore served as an exploratory reference population rather than a matched comparison cohort. As a result, differences observed between women with SLE and controls should be interpreted cautiously, particularly for outcomes strongly influenced by age and reproductive history.

## 5 Conclusion

This study provides an integrated view of reproductive health in women with SLE, encompassing pregnancy outcomes and long-term ovarian function. While most pregnancies resulted in favorable outcomes, maternal age and disease activity emerged as key determinants of complications, underscoring the importance of optimal disease control and individualized risk assessment. Longer disease duration was associated with early menopause, suggesting that cumulative disease burden may contribute to ovarian dysfunction and reproductive aging. Together, these findings support a shift from a pregnancy-centered approach to a life-course model of reproductive care in SLE, integrating routine assessment of menstrual health, fertility potential, and ovarian function to improve counseling and optimize reproductive health outcomes.

## 6 Acknowledgments

This work received support from Luis Aguilar, Alejandro León, and Jair García of the Laboratorio Nacional de Visualización Científica Avanzada. Authors would like to express their special acknowledgment to Fundación Proayuda Lupus Morelos A.C, Lupus MX, El despertar de la Mariposa, and Centro de Estudios Transdisciplinarios Athié-Calleja por los Derechos de las Personas con Lupus A.C., for their invaluable support.

## 7 Author contributions

GS-P: Conceptualization, Data curation, Formal analysis, Validation, Visualization, Writing – original draft, FB-G: Conceptualization, Data curation, Formal analysis, Validation, Visualization, Writing – original draft, MGMT: Resources, Writing – original draft, AM-G: Writing – original draft, AS: Resources, Writing – review & editing, NR-O: Resources, Writing – original draft, MBC: Resources, Writing – review & editing, SGGZ: Resources, Writing – review & editing, AP-A: Conceptualization, Resources, Writing – review & editing, LT-N: Conceptualization, Resources, Writing – review & editing, ET-V: Conceptualization, Resources, Writing – review & editing, DM: Conceptualization, Resources, Writing – review & editing, ALH-L: Conceptualization, Data curation, Formal analysis, Validation, Visualization, Writing – original draft, AM-R: Supervision, Funding acquisition, Writing – review & editing, DA-R: Conceptualization, Supervision, Writing – original draft, Writing – review & editing.

## 8 Statements and declarations

### 8.1 Ethical considerations

The study protocol was reviewed and approved by the Research Ethics Committee of the Instituto de Neurobiología at the Universidad Nacional Autónoma de México (UNAM, approval no. 093.H) on September 8, 2025.

### 8.2 Consent to participate

All participants provided informed consent prior to participating.

### 8.5 Data availability

All data is available upon request through the MexOMICS Consortium (https://redcap.link/nqsxtj8n). The complete analysis code is available through GitHub https://github.com/NeuroGenomicsMX/Reproductive_Health_SLE

