## Supplementary figures and images for "Reproductive health in Mexican women with systemic lupus erythematosus: pregnancy outcomes, menstrual irregularities and early menopause"

### SupplFig1.png

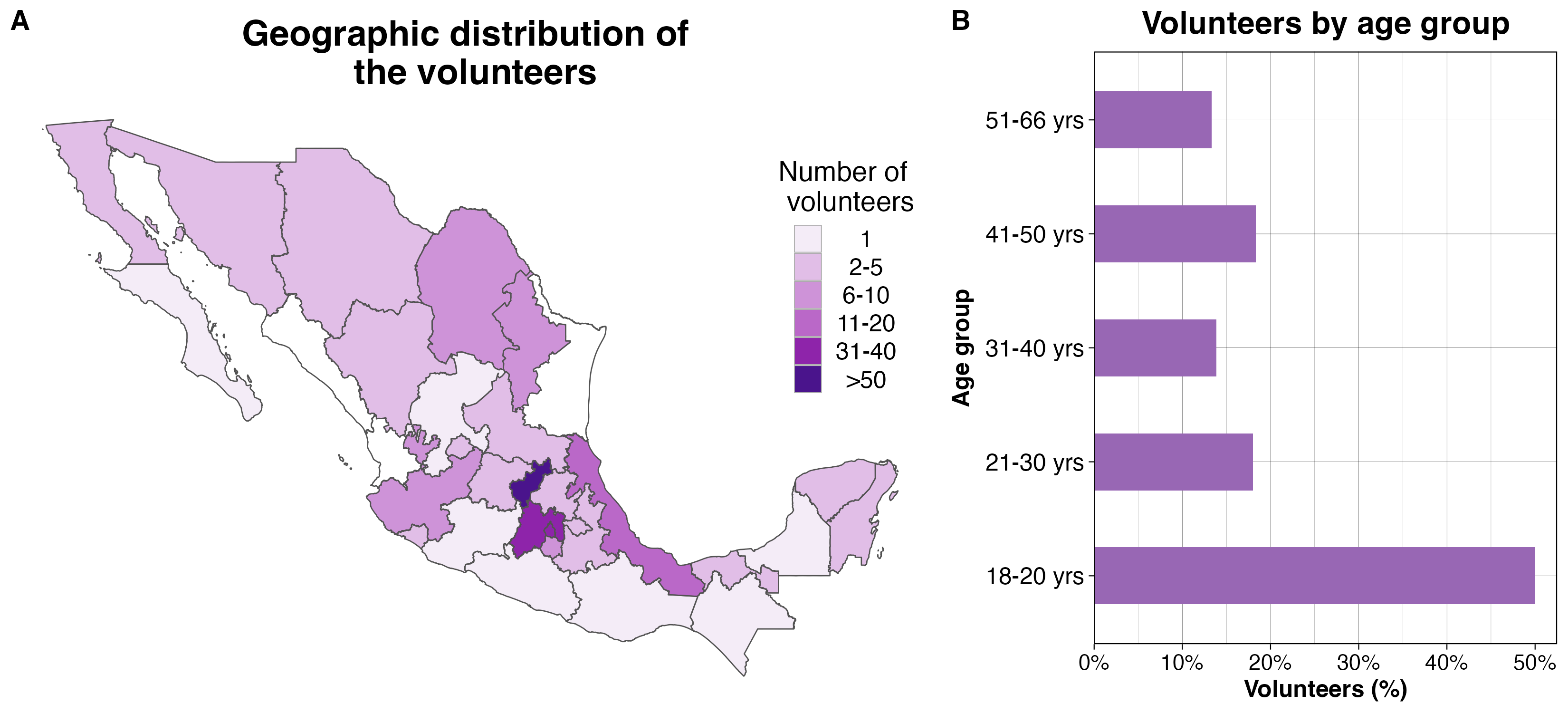

### SupplFig2.png

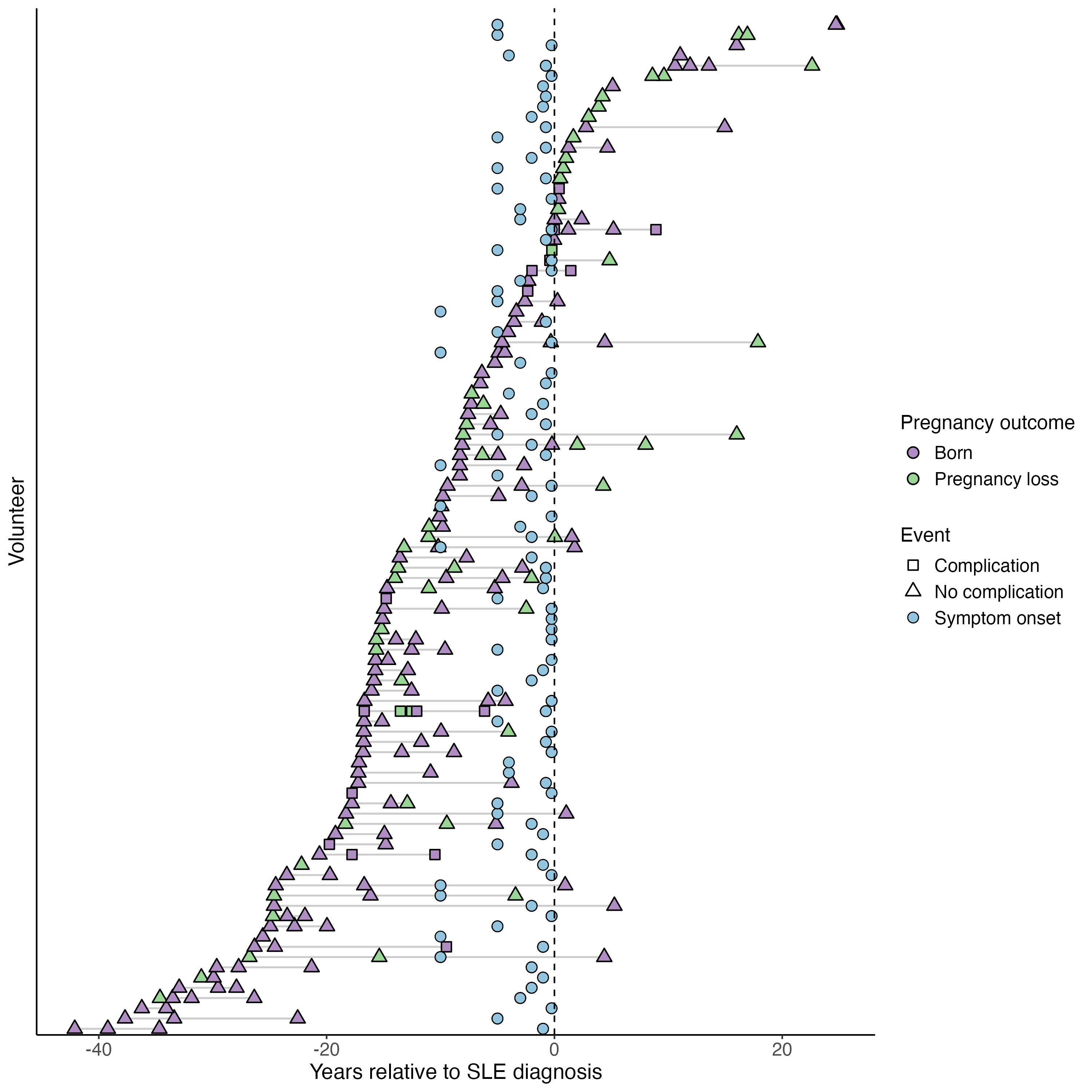
